# Estimating Under Reporting of Leprosy in Brazil using a Bayesian Approach

**DOI:** 10.1101/2020.05.22.20109900

**Authors:** Guilherme L. de Oliveira, Juliane F. Oliveira, Roberto F. S. Andrade, Joilda S. Nery, Julia M. Pescarini, Maria Y. Ichihara, Liam Smeeth, Elizabeth B. Brickley, Mauricio L. Barreto, Gerson O. Penna, Maria L. F. Penna, Mauro N. Sanchez

## Abstract

Leprosy remains an important health problem in Brazil - the country register the second largest number of new leprosy cases each year, accounting for 14% of the world’s new cases in 2019. Although there was increasing advances in leprosy surveillance worldwide, the true number of leprosy cases is expected to be much larger than the reported. Leprosy underreporting impair planning effective interventions and thoughful decisions about the distribution of financial and health resources. In this study, we estimated leprosy underreporting for each Brazilian microregion in order to guide effective interventions and resouce allocation to improve leprosy detection in the country. We extracted the number of new cases of leprosy from 2007 to 2015 and population and socioeconomic information from the 2010 Census for each Brazilian municipality and grouped data in microregions. We applied a Bayesian hierarchical model to obtain the best explicative model for leprosy underreporing using Grade 2 of leprosy-related disabilities as a proxy to explain the incidence rates. Then, we estimated the number of missing leprosy cases (underreported cases) and the corrected leprosy incidence rates for each Brazilian microrregion.

## Introduction

Leprosy is a Neglected Tropical Disease (NTD) caused by Mycobacterium leprosy and it is still concentrated in Low- and Middle-income countries and among individuals living under poor socioeconomic conditions (WHO 2019; Pescarini et al. 2018). The WHO Global Leprosy Strategy 2016-2020 reinforces the need to strenghten leprosy surveillance and health information systems for programme monitoring and evaluation and strengthen early detection through active case finding in leprosy endemic areas and among groups with increased risk (ie., household contacts of leprosy patients) (WHO 2016). Despite the recommendations, it was estimated that by 2020, there would be over 4 million leprosy cases undiagnosed and untreated worldwide (Smith et al. 2015).

Brazil is the second leading country in number of new leprosy cases, accounting for 14% of all new leprosy cases worldwide WHO 2019. Although large leprosy reductions took place in Brazil in the past decades, the proportion of cases with associated-leprosy disabilities have been increasing, which suggest that strategies to reinforce early detection and active case finding are still necessary (ref estudo descritivo). The Brazilian disease registration system is known to be reliable, but it still does not prevent small scale leprosy surveillance studies to find a substantial leprosy underreporting in both endemic and non-endemic regions of the country (Filho et al. 2017; Barreto et al. 2012). Leprosy underreporting can occur by many reasons, including the low capacity of health care services or health professionals to diagnose and register new cases of the disease, lack of specific leprosy programmes and policies, absence or poor national disease registries, or deficiencies of national or local leprosy programs (Filho et al. 2017; Barreto et al. 2012).

Previous studies have estimated leprosy underreporting and corrected existing estimates in specific regions of Brazil. A 2001 study conducted in a leprosy endemic municipality in the Northeast Brazil used a Bayesian spatial model to estimate leprosy underreporting among children and found correlation between underreporting and multibacillary forms of the disease (Souza et al. 2001). A recent study also used Spatial-temporal Bayesian models to estimate underreporting in an endemic State in the Northeast Brazil and correlated the presence of underreported areas (silent areas) to the higher presence of multibacillary cases and to the proportion of cases detected with Grade 2 (GD2) leprosy-related disabilities (Souza et al. 2018). Despite those efforts, no study to date provided national estimates of leprosy underreporting. Therefore, in this study we used a Bayesian hierarquical model aiming to estimate micro-regional level leprosy underreporting for Brazil in order to provide robust evidence that can be used to enhance national surveillance systems and to target specific policies to leprosy detection and control.

In this work we use a flexible Bayesian hierarchical model which leads to a complete predictive distribution for the true leprosy counts in Brazil. The severity of underreporting is estimated with basis on an available covariate that is potentially related to the reporting mechanisms. The considered Bayesian framework allows quantifying the uncertainty in correcting the underreporting.

## Materials and methods

### Data source

We used data from the Brazilian National Notifiable Diseases Information System (SINAN). We collected the total and new number of leprosy cases for each of the 558 mainland microregions of Brazil, from 2007 up to 2015. New cases are defined, by the Ministry of Health, as cases with no previous treatment (Brasil 2016). Confirmed cases of leprosy are all cases that presented one of the following conditions: (a) lesion(s) and/or area(s) of the skin with changes in thermal and/or painful and/or tactile sensibility; (b) peripheral nerve thickening associated with sensory and / or motor and / or autonomic changes; (c) presence of *Mycobacterium leprae bacilli*, confirmed by intradermal smear microscopy or skin biopsy. New cases of leprosy can be reported through an active detection, by performing epidemiological investigation of contacts and collective exams, such as surveys and campaigns (rapid serological tests), or by passive detection, when individuals spontaneously search health care systems.

The detection of the disease depends on the manifestation of symptoms in infected individuals. However, differences in the resistance of their immunological systems may contribute to the appearance of symptoms only after a long period after incubation of the bacterium, ranging from 2 up to 7 years (Brasil 2001). Additionally to such individual conditions, endemic levels and unfavorable socioeconomic factors, as well as access to public health services and a high rate of housing occupation, influence the pattern of the disease incidence in the country. Based on these factors, socioeconomic indicators and living conditions of the population in each microregion are considered to account for space variability of the disease incidence. Therefore, to conduct our analysis we collected the following information: (1) the proportion of examined household contacts with relation to the total registered household contacts (*x*_1_), obtainded from SINAN; (2) coverage of the cash transfer program Bolsa Familia in terms of the proportion of the population at risk that is assisted (*x*_2_); (3) coverage of the Family Health Program in terms of the proportion of registered population (*x*_3_); (4) average number of people per household (*x*_4_), obtained from the Atlas of Human Development in Brazil (2010); and (5) proportion of people living in urban areas (*x*_5_), obtained from the Brazilian Institute of Geography and Statistics - IBGE (2010 demographic census).

The evolution of symptoms will be reflected into disabilities, ranging from low, when patients have no problems in eyes, hands or feet, to high consequences, as visible deformations in hands, feet or eyes or even severe visual impairment. Therefore, new cases of patients presenting high levels of disabilities is an indirect indicator of diagnosis delay (Nobre et al. 2017), which leads to underreporting. Thus, in this work, the proportion of diagnosed new leprosy cases with Grade 2 of physical disability (*w*), collected from SINAN, was considered in the characterization of the underreporting mechanism throghout the Brazilian microregions.

### Data Curation

To peform our analises, we negleted missing informations. Complementary to this, from the total 311,970 new cases, with indentification of microregion, we excluded Fernando de Noronha from the analysis, once it is an island that stays more than 500 km from the mainland Brazil and might generate problems for the spatial analysis. Therefore, for the spatial analysis, we considered 557 mainland Brazilian microregions. After that, it remained 311,968 new cases from the period 2007 a 2015.

To obtain the numerator to calculate the proportion of diagnosed new leprosy cases with GD2, we exclude from the analysis the null and ”not evaluated” values from the variable type of diagnosis. They represented 10.35% of the total number registered. Therefore, 279,719 had information about the type of diagnosis.

The proportion of household examined contacts was obtained as the ratio between the number of examined contacts and the number of identified contacts in each microregion. We excluded the null values and the cases with less than 43 contacts in those variables. We obtained a total of 267,160 with information.

### Statistical model for underreported leprosy data in Brazil

In disease surveillance and epidemiological studies, two main statistical approaches have been considered to deal with underreport counts. Inference under the first one is based on a censored Poisson likelihood function, allowing the estimation of both the disease rate and the probability of the observed count being underreported in each area (Bailey et al. 2005; Oliveira et al., 2017). The second and potentially more flexible approach relies on the specification of a hierarchical Poisson model from which is possible to estimate both the disease rates and the proportion of reported cases in each area (Stoner et al. 2019; Dvorzak and Wagner 2015; Shaweno et al. 2017; Stamey et al. 2006; Whittemore and Gong 1991; Papadopoulos and Silva 2012). Based on the sample information we have available, we consider the Bayesian framework proposed by Stoner et al. 2019 to fit the Brazilian leprosy data.

Under such a framework, in each microregion *i* (for *i* = 1,…, 557), the reported (observed) count *Y_i_* is modeled as a Binomial random variable, where the number of trials is an unobserved Poisson variable *T_i_* corresponding to the true number of cases that has been incompletely recorded. The true count generating process is modeled through the mean of the Poisson variable, denoted by *μ_i_*, and the reporting mechanism is modeled through the Binomial probability, denoted by *ϵ_i_*. Then, the basic structure for the hierarchical model is:

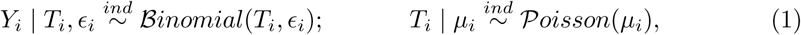

for *i* = 1,…, 557. Since *T_i_* is not observed, statistical inference is based on the marginal distribution of *Y_i_* obtained from the joint model given in equation (1), which is

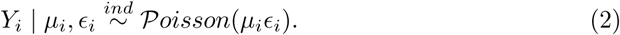

We assume that the mean expected new cases of leprosy, *μ_i_*, depends on the five socioeconomic covariates *x*_1_ to *x*_5_ previously described, such that

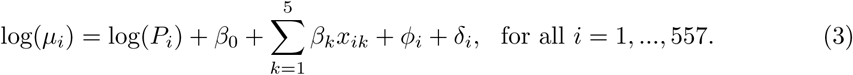

We also include the logarithm of the total exposure population, *log*(*P_i_*), as an offset, so that parameter *μ_i_* can be interpreted as the leprosy incidence rate at area *i*. Additionally, in order to capture any residual variation in the leprosy, we include a spatially structured random effect *ϕ_i_* and a local unstructured random effect *δ_i_* in the log-linear predictor of *μ_i_*.

To model *ϵ_i_*, the probability of reporting a new leprosy case at each area *i*, we made use of the covariate *w*: the percentage of diagnosed new leprosy cases with GD2 of physical disability, such that

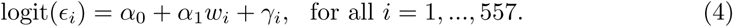

As previously discussed, covariate *w* acts as a *proxy* of the appropriate variable that accounts for notification efficiency of leprosy new cases in each microregion. An unstructured random effect *γ_i_* is included in the logistic regression model for *ϵ_i_* in order to account for potential effects of unobserved covariates that may influence the detection of leprosy.

In the statistical literature, the joint model defined by equations (2), (3) and (4) is called by the Poisson-Logistic (or Pogit) model. It worth noticing that the Pogit model provides a straightforward predictive analysis for the proportion of leprosy new cases that were not observed, denoted by *Z_i_* = *T_i_* − *Y_i_*. Given and *ϵ_i_*, *Z_i_* can be predicted from the distribution 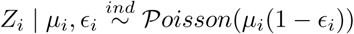 for all *i*.

Despite its appealing features, it is well-known in the literature that the Pogit model suffers with the lack of identifiability. This occurs because only the product *η_i_* = *μ_i_ϵ_i_* is identified from the observed data since any other parameter combination, say 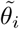 and 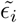, such that 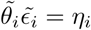 yields the same likelihood function. In practice, that concept means that additional information must be introduced in the model in order to distinguish between parameters *μ_i_* and *ϵ_i_*, thus conducting to a meaningful inference about the true count *T_i_*. Such extra information can be provided by validation datasets, active search surveys and experts’ opinion. The source of information to be used will depend on which one is available for the specific practical situation one is dealing with.

To overcome the identifiability issue when fitting the Pogit model to the Brazilian leprosy count data, we follow the approach of Stoner et al. 2019. Whenever the regression models of *μ_i_* and *ϵ_i_* do not share any common covariate, the lack of identifiability of the Pogit model relies on the confounding between the two intercepts *α*_0_ and *β*_0_. Then, special attention must be given to their prior distributions. As discussed in Stoner et al. 2019, by taking *x*_1_ to *x*_5_ and *w* as centered covariates it provides that *β*_0_ and *α*_0_ are, respectively, interpreted as the mean reported number of leprosy new cases (on the log scale) and the mean reporting rate (on the logistic scale) when the covariates are at their centering value. In this context, the appealing interpretation of *α*_0_ and *β*_0_ can be appropriately used to elicit an informative prior distribution for one of them, thus providing an identifiable Pogit model.

We found that there is no study seeking to estimate the detection rate of leprosy over the country between the period of 2007 to 2015. If such a study was available, we could center the covariate *w* with respect to its observed mean and then elicit an appropriate informative prior for parameter *α*_0_, which, in this case, could be interpreted as the overall mean reporting rate. In the work by Cunha et al. 2015, the authors present results of an active search survey, performed in 2012, in some municipalities of the state of Amazon, Northern of Brazil. Based on their findings, the microregion *Rio Preto da Eva* (RPE), composed by the municipalities *Rio Preto da Eva* and *Presidente Figueiredo*, showed reporting rates of approximately 91% when comparing the total cases registered by the local health centers. In particular, the microregion of RPE is located in the metropolitan region of *Manaus*, capital of the State of Amazonas, and it is one of the most developed regions in northwest of Brazil.

As far as we know, this is the most trustful available study about the level of leprosy underreporting in a Brazilian microregion. Therefore, we rely on the information provided for this microregion to define an informative prior distribution for parameter *α*_0_. To do so, we center the covariate *w* with respect to its observed value for Itacoatiara so that the parameter *α*_0_, on the logarithmic scale, represents the average level of notification of leprosy cases in such microregion. Moreover, we assume that the information for the level of notification for the period from 2007 to 2015 can be represented by findings of Cunha et al. 2015 for year 2012. We then specified a *N*(2.5, 0.3) as the prior distribution for the parameter, which provides an *a priori* average level of reporting of approximately 91% for this microregion, on the logistic scale. The choice for the variance term was guided by the sensitivity studies presented by Stoner et al. 2019. They suggest that the model is robust in terms of quantifying uncertainty, as long as the practitioner specifies a prior for *α*_0_ that is informative without the need of being a degenerated one (a prior with null variance). Through an a priori predictive analysis in our application, the variation expressed by the prior distribution *N*(2.5,0.3) provides values for *ϵ* in the microregion RPE with high concentration (about 90%) between 88.2% and 95.1%, reflecting well our belief regarding the average level of reporting used as a reference for the period.

We now provide the prior distribution for parameter *β*_0_. As an informative prior was already elicited for the other model intercept, *α*_0_, there is no need for doing the same in relation to *β*_0_. Nevertheless, to avoid the generation of unrealistic values for the leprosy incidence rate (especially quite elevated values), we also follow the approach proposed in Stoner et al. 2019. Each covariate *x*_1_ to *x*_5_ was centered with relation to its observed mean such that is interpreted as the mean reported number of leprosy cases, on the log scale. Then, we assume a prior *N*(−8,1) for *β*_0_ to represent our belief that is not plausible a very high value (such as over 1 million) for the total number of new leprosy cases. For each of the remaining regression coeficients *α*_1_ and *β*_1_, …, *β*_5_ we elicit a relatively noninformative Normal prior distribution *N*(0,10^2^).

The model specification is completed with the following prior distributions for the random effects and their precision terms. As it is usual in the literature, the *prior* distribution of *ϕ_i_*, with precision parameter *ν*, is represented by an intrinsic conditional autoregressive (iCAR) model (Besag et al. 1991). Here, a neighbor of an area *i* was defined as any *i*′ ≠ *i* sharing a geographical boundary with *i*. Both the unstructured effect *γ_i_* and the local effect *δ_i_* are assumed to have a Normal *prior* distribution 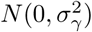, 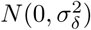 and 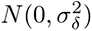, respectively. For each of the precision parameters *v*, 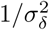 and 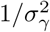 we elicited a Gamma distribution *G*(1,1).

The model was implemented using package NIMBLE from software R (R Core Team, 2015). For the MCMC scheme two chains were used, each ran for a total of 600K iterations. After discarding 200K iterations as burn-in and using a lag of 200, the potential scale reduction factor (PSRF) proposed by Brooks and Gelman (1998) was computed as less than 1.05 for all regression coefficients and precision parameters, indicating convergence to the target posterior distribution.

To summarise, we applied the Bayesian model to correct the underreporting of leprosy new cases, from which we estimate the posterior mean (Mean), the posterior standard deviation (SD) and the 95% highest posterior density (HPD) intervals (95%-HPD) for all regression parameters defined on Equations (3) and (4) (Table 1). We also provide the incidence rate ratio (IRR) for Poisson parameters and the odds ratio (OR) for logistic parameters with their respective 95% HPD intervals (95%-HPD).

**Table 1.** Posterior summaries for the regression effects *β* and *α* and the model variance parameters; Brazilian leprosy data 2007-2015.

| Poisson | Covariate Name | Mean | SD | 95%-HPD | IRR (95%-HPD) |
| --- | --- | --- | --- | --- | --- |
| $\beta_0$ | mean reported number of leprosy cases (log scale) | -8.910 | 0.034 | (-8.977, -8.844) | - |
| $\beta_1$ | proportion of household contacts examined | 0.739 | 0.294 | (0.177, 1.301) | 2.094 (1.081, 3.502) |
| $\beta_2$ | coverage of program Bolsa Família | 1.377 | 0.611 | (0.222, 2.619) | 3.963 (0.710, 1.071) |
| $\beta_3$ | coverage of the Family Health Program | 0.034 | 0.178 | (-0.314, 0.372) | 1.034 (0.722, 1.434) |
| $\beta_4$ | average number of people per household | -0.364 | 0.120 | (-0.607, -0.135) | 0.695 (0.539, 0.867) |
| $\beta_5$ | proportion of people living in urban areas | 0.682 | 0.232 | (0.253, 1.146) | 1.978 (1.197, 3.002) |
| $\nu$ | spatial effect precision parameter | 0.975 | 0.095 | (0.798, 1.162) | - |
| $\sigma_\delta$ | unstructured effect variance parameter | 0.085 | 0.015 | (0.057, 0.115) | - |
| Logistic | Covariate Name | Mean | SD | 95%-HPD | OR (95%-HPD) |
| $\alpha_0$ | mean reporting rate (logistic scale) | 2.548 | 0.330 | (1.8306, 3.1798) | - |
| $\alpha_1$ | proportion of diagnosed new leprosy cases with GD2 | -0.061 | 0.022 | (-9.7326, -1.9471) | 0.941 (0.903, 0.982) |
| $\sigma_\gamma$ | unstructured effect variance parameter | 0.608 | 0.324 | (0.102, 1.258) | - |

For each model coefficient, we provide the IRR (or OR) and its 95% HPD interval. When such interval contains the value 1 it can be considered that the covariate effect is not significant. The same if the 95% HPD interval for the coefficient contains the value zero. The IRR indicates the effect in the mean incidence when a unity is increased in the explanatory variable.

## 1 Ethic statements

This study was approved by the ethics committees of the University of Brasília (UnB) (1.822.125), Instituto Gonçalo Muniz - Fiocruz (1.612.302), and London School of Hygiene Tropical Medicine (10580 – 1).

## Results

From January 2007 to December 2015, a total of 312,114 new cases of leprosy were registered in SINAN, of which 7.6% of had GD2 at diagnosis. The highest mean incidences of leprosy in the period are concentrated in the North (42.95/100,000 inhabitants), Northeast (26.53/100,000 inhabitants) and Central-west (40.87/100,000 inhabitants of Brazil. In the Central-West region, 46% were detected in Mato Grosso. Froma 2007 to 2015, all Brazilian states showed a decreasing number of leprosy cases over time, except for Mato Grosso do Sul, which presented a 6% increase in leprosy incidence during this period. The states with the highest percentage of cases of GD2 include Tocantins (12.33%), Maranhão (14.76%), Mato Grosso (12.06%); São Paulo (8.37%); and Rio Grande do Sul (6.21%).

We found a clear spatial variability in the observed mean incidence rate across Brazilian microregions over 2007 and 2015, which cam be partially attributed to measured covariates (Fig. 1a). A spatial pattern can also be noted in the proportion of new leprosy cases of GD2 of physical disability (Fig. 1b). With exception for the extreme South of Brazil, regions with lower incidence rates tends to present a higher proportion of new leprosy cases of GD2.

**Fig 1.**
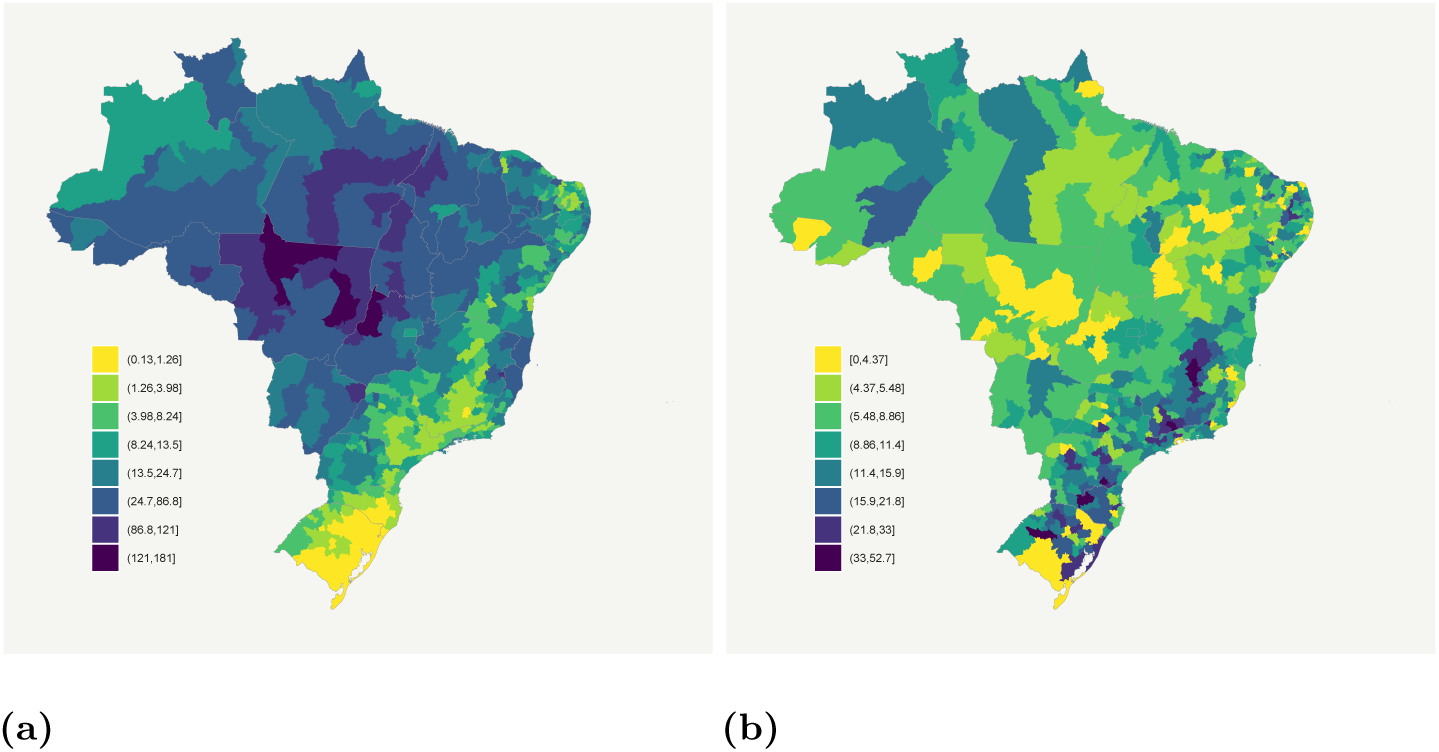
Brazilian microregions: (a) Observed mean leprosy incidence rate per 100,000 inhabitants over the years 2007 to 2015; (b) Proportion of diagnoses of new leprosy cases with GD2 of physical disability over the years 2007 to 2015.

The incidence rates displayed in Fig. 1a are likely underestimated due to the amount of new leprosy cases that go unreported.

In Fig. 2 we present an exploratory analysis of the relation between the observed leprosy incidence and the variables to be considered in the model.

**Fig 2.**
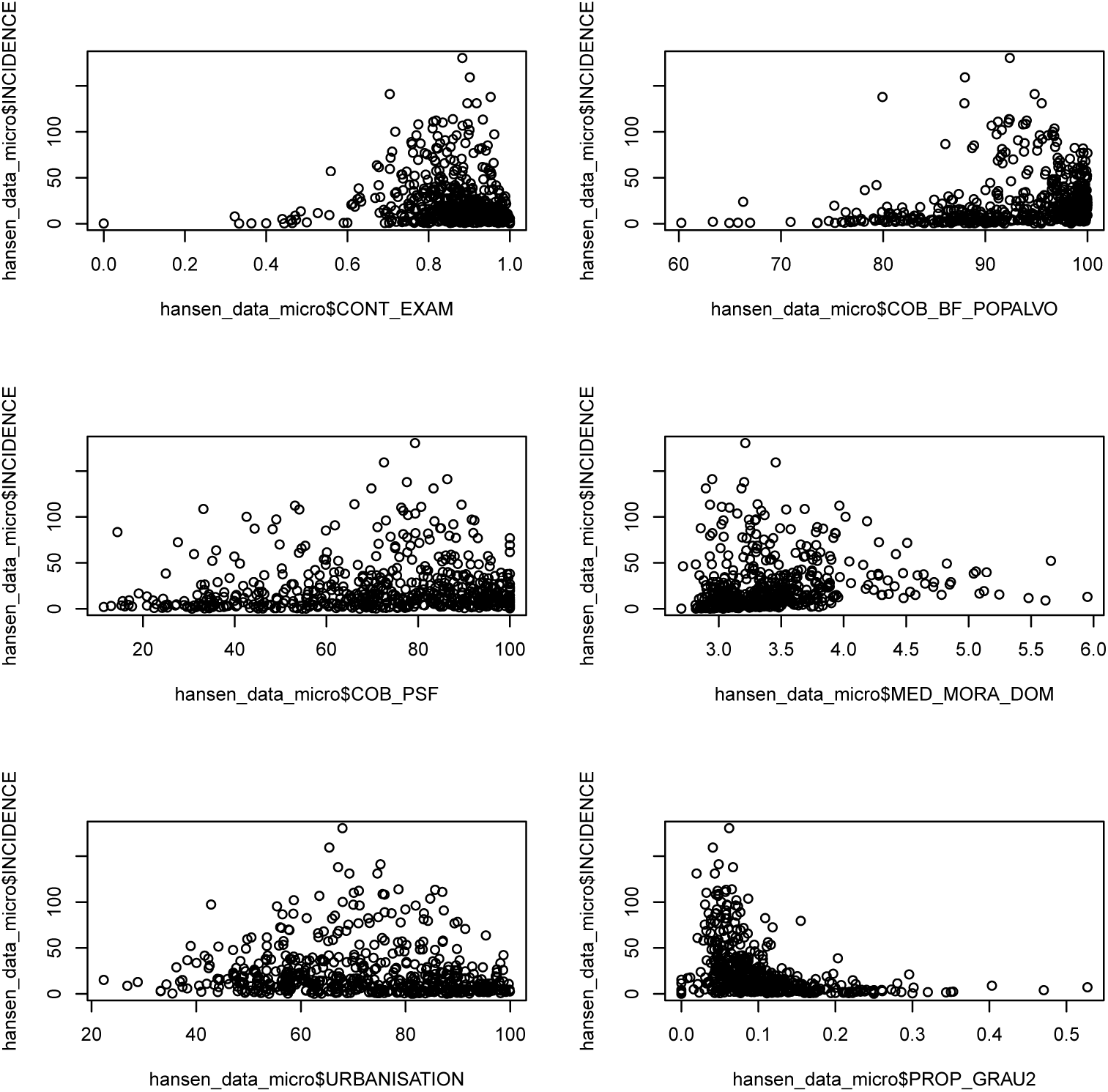
Relations between the observed incidence and the variables to be considered in the model.

According to the 95%-HPD intervals presented in Table 1, the result indicates that only *Family Health Program coverage* is does not explain the incidence of leprosy new cases in the period, since the interval for parameter *β*_3_ contains the value zero. The *average number of people per household* shows to have a negative impact in the leprosy incidence (IRR=0.70). Therefore, keeping all other variables in the same value, an unit increase in the *average number of people per household* makes the average leprosy incidence ratio be around 44% smaller. The negative effect can be explained by both the variability of household crowding and the presence of microregions with extreme values and low incidence 2.

The effects of the *proportion of household contacts examined* and the *proportion of people living in urban areas* are similar. Individually, the average leprosy incidence is expected to double when one of the variables is increased by one unit. Using a similar analysis, we note that the *coverage of program Bolsa Família* present the largest impact on leprosy incidence rates (IRR=3.96), which means that for 100 percent increase in *coverage of program Bolsa Família*, the mean incidence of leprosy increases about 4 times.

Based on the logist model, the *proportion of new leprosy cases with GD2 at diagnosis* is a significant construct to explain the leprosy reporting process in the Brazilian microregions. As the proportion of cases with GD2 of physical disability increases, the reporting probability decreases. We found that an unit increase in this construct tends to make the reporting probability to be around 6% smaller (or to be multiplied by 0.941).

It is estimated that, in average, 29,649 (95%-HPD = (14,300; 50,051)) new cases of leprosy were not reported from the years of 2007 to 2015, giving an overall leprosy detection rate in Brazil of approximately 91.32% for the period (Table 2). The posterior estimate of the probabilities of leprosy undereporting, explained by the covariate of GD2, show that the South and Southwest regions of Brazil present the lowest probability of reporting a case - 85% and 87%, respectively (Fig. 3). Nevertheless, some microregions in the South and South-east States reported 61% of the expected number of leprosy cases. Amazonas, Roraima and Amapá (all in the North region) detected 85.22%, 87.96%, 87.78% of the true number of leprosy cases, while the Federal District (Central-west) detected only 83.71% of the true/expected number of leprosy cases (Table 2).

**Fig 3.**
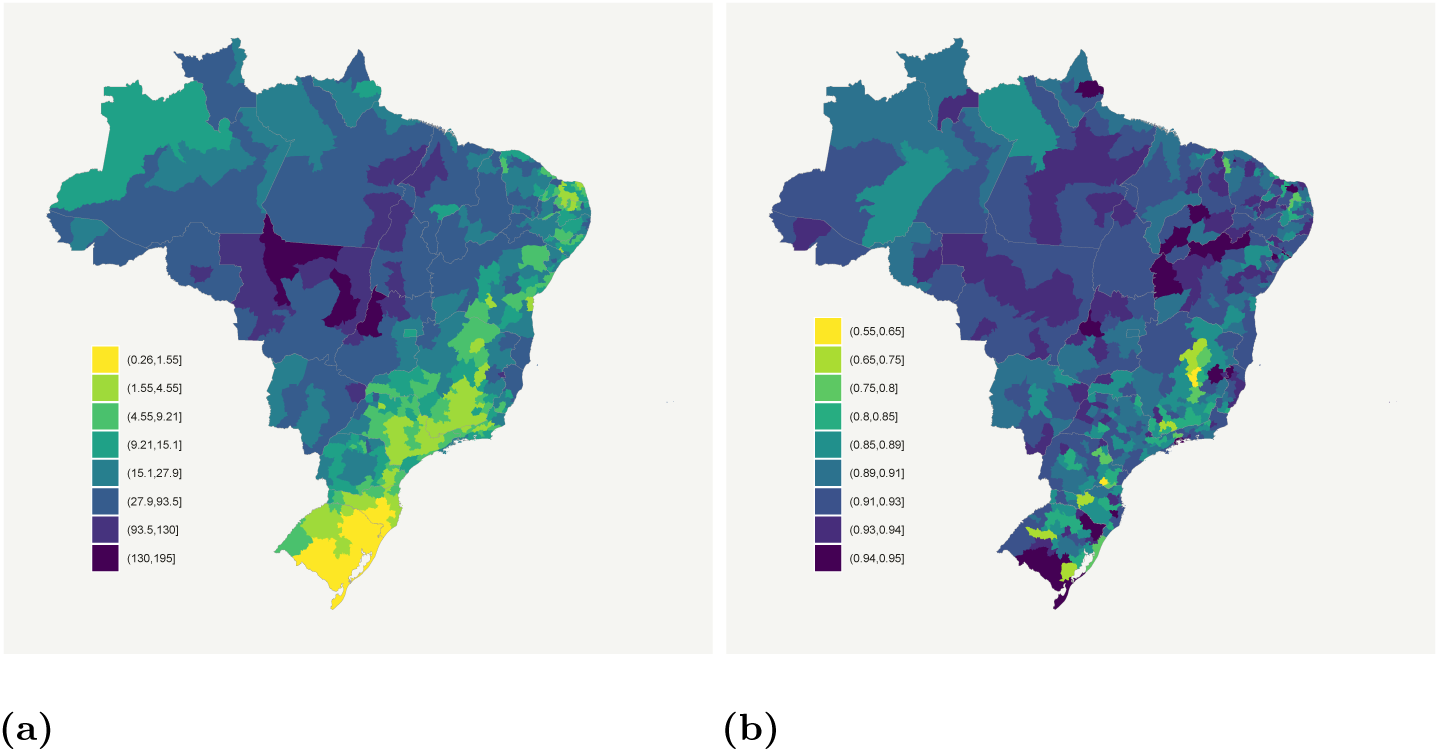
Brazilian microregions: (a) Posterior mean leprosy incidence rate per 100,000 inhabitants over the years 2007 to 2015 corrected by underreporting; (b) Posterior estimates for reporting probabilities, for each Brazilian microregions.

**Table 2.** Observed and total (estimated) number of leprosy cases and overall leprosy detection rate for each Brazilian State for the period 2007-2015.

| Brazilian States | Observed<br>number<br>of cases (Y) | Unobserved<br>estimated<br>number<br>of cases (Z ) | Total corrected<br>estimated<br>number<br>of cases (T) | Overall leprosy<br>detection rate<br>($\frac{Y}{T}\%$) |
| --- | --- | --- | --- | --- |
| Rondônia | 7899 | 710 | 8609 | 91.75 |
| Acre | 1878 | 165 | 2043 | 91.92 |
| Amazonas | 5955 | 697 | 6652 | 89.52 |
| Roraima | 1235 | 130 | 1365 | 90.48 |
| Pará | 34041 | 2997 | 37038 | 91.91 |
| Amapá | 1293 | 132 | 1425 | 90.74 |
| Tocantins | 9589 | 844 | 10433 | 91.91 |
| Maranhão | 35533 | 3244 | 38777 | 91.63 |
| Piauí | 11446 | 950 | 12396 | 92.34 |
| Ceará | 19278 | 1795 | 21073 | 91.49 |
| Rio Grande<br>do Norte | 2532 | 268 | 2800 | 90.42 |
| Paraíba | 6093 | 588 | 6681 | 91.20 |
| Pernambuco | 24518 | 1992 | 26510 | 92.49 |
| Alagoas | 3475 | 349 | 3842 | 90.87 |
| Sergipe | 3897 | 374 | 4271 | 91.24 |
| Bahia | 23968 | 2081 | 26049 | 92.01 |
| Minas Gerais | 14117 | 1842 | 15959 | 88.46 |
| Espírito Santo | 8291 | 684 | 8975 | 92.38 |
| Rio de Janeiro | 15117 | 1464 | 16581 | 91.17 |
| São Paulo | 15835 | 1898 | 17733 | 89.30 |
| Paraná | 9343 | 1204 | 10547 | 88.58 |
| Santa Catarina | 1739 | 236 | 1975 | 88.05 |
| Rio Grande<br>do Sul | 1331 | 195 | 1526 | 87.22 |
| Mato Grosso<br>do Sul | 6486 | 699 | 7185 | 90.27 |
| Mato Grosso | 24902 | 2044 | 26946 | 92.41 |
| Goiás | 20183 | 1798 | 21981 | 91.82 |
| Distrito Federal | 1994 | 269 | 2263 | 88.11 |
| Total | 311968 | 29649 | 341617 | 91.32 |

## Discussion

This study demonstrated a clear spatial variability in the mean incidence rates across micro regions, which is related to spatial covariates and is consistent with the spatial variability of socioeconomic indicators in Brazil, as well as the quality and access to health care systems and exposure to high risk factors for leprosy. In other words, individuals with poor socioeconomic status or living under deprived conditions continue to be more likely to be affected by leprosy and leprosy associated disabilities. Proportion of new cases with G2D also showed a defined spatial pattern.

The reporting probability decreases with increasing proportions of cases with G2D. This may indicate late diagnosis and potentially missed opportunities to start treatment as one or more contacts with the public health system might have occurred. Alternatively, it can be a cultural issue - according to Penna et al. 2009, “the diagnosis of skin diseases depends on the cultural importance given to skin lesions, as well as health-seeking habits among the population.” Yet, this phenomenon may be explained by lack or training from the healthcare staff, as they are not seeing leprosy frequently as these areas are not of high endemicity. The spatial pattern of leprosy underreporting and, therefore, the correction of the true leprosy incidence follows the one for G2D frequency. This reinforces the hypothesis that health care staff does not have leprosy diagnoais as part of their routine, and the population does not look for care on a timely manner, as G2D is considered a marker for late diagnosis. All these hypotheses deserve further evaluation and should be the focus of future studies.

The variables found to be associated with incidence of leprosy in our study are in line with the literature. Average number of people per household (protective effect), proportion of household contacts examined and coverage of the Bolsa Familia Program (risk factors). Nery et al. 2019 shows that increased levels of deprivation and less schooling were associated with higher levels of new case detection. Considering that Bolsa Família is a conditional cash transfer program targeting individuals, especially families with a woman as the head of household, on social vulnerability situation, our finding is plausible. In this case, we are considering presence and coverage of this cash transfer program as a marker for higher levels of social and economic deprivation.

The proportion of examined contacts is a possible marker of the effort the leprosy local control program on contact examination. For the actual incidence rate to decline because of these actions, a longer period of follow up would be needed. Ultimately, obtaining reduction in incidence of disease is the goal, as outlined in all plans leading to leprosy control WHO 2016.

Our estimates show that almost 30,000 cases were missed by the surveillance system during the 5 years of study. This is an important figure when one considers the need identify cases, ideally early in the course of disease progression, to break the transmission chain and prevent the development of physical disability. The recommendation is that special attention should be given to areas where the need for correction for underreporting was identified to be higher. This way the country can address this severe public health issue in a more effective manner, paving the way for fulfilling the national and international agreements regarding leprosy control.

## Data Availability

All data are available as supplementary material.

## Acknowledgements

This study was funded by The Medical Research Council (MR/N017250/1), CONFAP/ESRC/MRC/BBSRC/CNPq/FAPDF - Doenças Negligenciadas (FAP-DF to Gerson O. Penna, (Number 193.000.008/2016). Wellcome Trust (202912/B/16/Z), the funders of the study had no role in the design, data collection, analysis, interpretation, or writing the article. JFO was supported by the Center of Data and Knowledge Integration for Health (CIDACS) through the Zika Platform - a long-term surveillance platform for Zika virus and microcephaly (Unified Health System (SUS), Brazilian Ministry of Health).

## Supporting information

**S1 File. Brazilian microreagion leprosy data used in the manuscript.**

## Notes

### Competing Interest Statement

The authors have declared no competing interest.

### Funding Statement

This study was funded by The Medical Research Council (MR/N017250/1), CONFAP/ESRC/MRC/BBSRC/CNPq/FAPDF - Doencas Negligenciadas (FAP-DF to Gerson O. Penna, (Number 193.000.008/2016). Wellcome Trust (202912/B/16/Z), the funders of the study had no role in the design, data collection, analysis, interpretation, or writing the article. JFO was supported by the Center of Data and Knowledge Integration for Health (CIDACS) through the Zika Platform - a long-term surveillance platform for Zika virus and microcephaly (Unified Health System (SUS), Brazilian Ministry of Health).

### Author Declarations

This study was approved by the ethics committees of the University of Brasilia (UnB) (1.822.125), Instituto Goncalo Muniz - Fiocruz (1.612.302), and London School of Hygiene & Tropical Medicine (10580-1).

